# Perceptual discrimination of complex objects: APOE e4 gene-dose effects in mid-life

**DOI:** 10.1101/2024.10.18.24315682

**Authors:** Claire Lancaster, Sam Berens, Jessica Daly, Jennifer Rusted, Chris M. Bird

## Abstract

**INTRODUCTION:** Complex perceptual discrimination is supported by tau-vulnerable regions of the medial temporal lobe; notably the perirhinal cortex. This research tests whether there is a gene-dose effect of Apolipoprotein e4 on perceptual discrimination in mid-life.

**METHODS:** 313 adults (45-65 years; grouped by APOE e4 gene-dose (142 APOE33, 135 APOE34, 36 APOE44)) completed a Greebles ‘odd-one-out’ task.

**RESULTS:** APOE44 carriers were significantly less accurate in their perceptual judgements than APOE33 and APOE34 individuals. There was a significant Age x APOE e4 gene-dose interaction in response speed, with the slope of age-related slowing increasing stepwise with the number of e4 alleles carried. Estimates suggest high-risk individuals are quicker than the APOE33 control group until age 54, but slower thereafter.

**DISCUSSION:** Perceptual disadvantages specific to APOE44 individuals suggest this high-risk group show compromised MTL-function by mid-life, potentially through accelerated tau-aggregation. Task performance in APOE34 carriers is relatively preserved across the studied age range.

**Research in Context:**

1. **Systematic review:** A review of the literature (e.g., PubMed; Google Scholar databases) revealed perceptual discrimination is impaired in the mild to moderate stages of Alzheimer’s Disease. No prior study, however, tests for an effect of APOE e4 gene-dose in healthy adulthood.
2. **Interpretation:** Homozygous carriers of an APOE e4 genetic risk variant show perceptual disadvantages in mid-life. This may possibly reflect compromised MTL-function in this group, consistent with the theory perceptual discrimination will be vulnerable to early transentorhinal tau-accumulation. There is limited evidence of impairment in individuals carrying one copy of APOE e4 in mid-life.
3. **Future directions:** Whether transentorhinal and hippocampal tau mediates the relationship between APOE e4 gene-dose and perceptual discrimination remains to be established. Future research may also explore the utility of perceptual discrimination tasks in clinical trials targeting the preclinical stages of Alzheimer’s Disease.

## 1. Introduction

To progress interventions targeting the early, clinically ‘silent’ stages of Alzheimer’s Disease (AD), the field needs sensitive and scalable tools for identifying individuals at risk of future neurodegeneration. Behavioural markers may be best identified by considering the topographical staging of AD neuropathology and identifying the cognitive functions supported by brain regions with early vulnerability. Such markers must be capable of detecting subtle differences in outwardly healthy individuals, as well as predicting further AD-biomarker progression, and later conversion to dementia [1:3]. This requires a step away from standardised neuropsychological assessment which dominates dementia research to date [4:6].

Tests of perceptual discrimination, specifically those requiring participants to distinguish between representations with a large proportion of overlapping visual features, are a candidate cognitive marker of AD-risk. Such measures are sensitive to disadvantages in individuals with Mild Cognitive Impairment and AD [7:8]. Indeed, older adults without a dementia diagnosis who have been classified as ‘high-risk’ based on their neuropsychological test scores show impaired ability to distinguish between ‘Greebles’ – abstract 3D-presented objects [9:10]. Perceptual discrimination is supported by regions of the medial temporal lobe (MTL), including the anterolateral entorhinal cortex (alERc) and perirhinal cortex (PRc) [11:14], with the PRc believed to act as an interface between the ventral visual processing stream, higher-level perceptual, and memory systems [15:16]. Given neurofibrillary tau pathology initiates in the PRc, before spreading into the entorhinal and hippocampal regions of the MTL [17:18], perceptual discrimination tasks may be a valuable indicator of ‘preclinical’ AD. However, there is no evidence to date establishing the utility of such measures for detecting differences in mid-age adults with early tau pathology.

The Apolipoprotein (APOE) e4 variant is the leading genetic risk factor for sporadic AD [19], yet there remains considerable debate as to when in the lifespan detrimental genotype effects emerge and via which mechanisms [4, 20:22]. Consistent with APOE e4 promoting earlier toxic gain of neurofibrillary tau [23:25], complex perceptual discrimination tasks are expected to be sensitive to genotype disadvantages by mid-life. However, to date there has been limited investigation as to whether APOE e4 carriers show impaired perceptual discrimination, with equivalent performance reported between APOE e4 carriers and an APOE33 control group in early adulthood, including across multiple stimulus modalities [26]. Middle-aged individuals with a family history of dementia, however, show disadvantages on a ‘Greebles’ odd-one-out task [27], supporting a genetic link.

Here, we tested if APOE e4 carriers show differences in simple and complex perceptual discrimination in mid-life, using a remote online version of the ‘Greebles’ task [28]. In total, 313 individuals completed this task; inclusion of a higher proportion of APOE34 and APOE44 individuals than typically found in the population [29] allowed novel exploration of gene-dose effects. The lifetime risk of dementia is substantially higher in APOE44, relative to APOE34, with 60% of homozygous individuals receiving a diagnosis of AD by 85 years [30].

Furthermore, a recent study reports AD neuropathology (including tau) increases dramatically in APOE44 individuals from age 50, with dementia symptoms emerging earlier (mean age 65.6 years) in this high-risk group [31]. As such, we predicted a stepwise disadvantage in perceptual discrimination with the number of APOE e4 alleles carried. Such a difference would be consistent with enhanced early vulnerability to tau in this group.

## 2. Methods

Hypotheses, experimental design, and statistical procedures were pre-registered on the Open Science Framework in advance (https://doi.org/10.17605/OSF.IO/F67XJ).

### 2.1 Participants

Three-hundred and thirteen cognitively healthy middle-aged adults (45-65 years old) were recruited from NIHR BioResource as part of a larger online study: The APOE Memory Bank. Exclusion criteria included any self-reported dementia, neurological, or current psychiatric diagnoses. In addition, APOE e2 carriers were excluded due to the reported protective effects of this less common APOE variant against Alzheimer’s disease [32,33]. Participants were selected with a bias towards APOE e4 carriers, with 5-year age band and gender matched by APOE status (Table 1). Most participants (97.73%) self-reported as white or white British.

**Table 1.** Demographic characteristics of participants grouped by APOE e4 carrier status and gene dose. Note. Immediate family history refers to having a parent or sibling with a dementia diagnosis.

|  | APOE e4 status |  | APOE e4 gene dose |  |
| --- | --- | --- | --- | --- |
|  | APOE33 | APOE e4+ | APOE34 | APOE44 |
| *n* | 142 | 171 | 135 | 36 |
| **Gender (%F)** | 52.81 | 50.29 | 51.11 | 47.22 |
| **Age Band (years, n)** |  |  |  |  |
| **45-49** | 32 | 25 | 24 | 1 |
| **50-54** | 30 | 41 | 36 | 5 |
| **55-59** | 34 | 45 | 32 | 13 |
| **60-65** | 45 | 60 | 43 | 17 |
| **Immediate Family History of Dementia (%)** | 18.31 | 25.15 | 25.93 | 22.22 |
| **Degree-level education (%)** | 53.52 | 55.56 | 54.81 | 58.33 |

All participants gave informed electronic consent, including acknowledgement that participants would not receive feedback on their APOE status. The study was approved by the University of Sussex Science & Technology Research Ethics Committee (ER/CLL6/2) and was completed in accordance with the ethical standards outlined in the 1964 Declaration of Helsinki (and its later amendments).

### 2.2 “Odd-one-out” task

The ‘odd-one-out’ task is a forced-choice measure of perceptual discrimination. In each trial, participants were shown three computer-generated, fictional objects (Greebles – http://www.tarrlab.org) from marginally different viewpoints. Two of these objects are the same and one is different. Participants were asked to select the ‘odd-one-out’, indicating their choice by clicking on a button on the screen. There is no limit on how long participants can take to respond, however, participants were instructed to respond as accurately and as quickly as possible.

In the low ambiguity condition (24 trials), the presented Greebles [28] share few perceptual features (e.g., the position of an appendage) and hence discriminating between them places relatively low demand on perceptual processes. In the high ambiguity condition (36 trials), exemplar Greebles are matched on several features (e.g., body shape, basic shape, and position of appendages). As such, discrimination requires a conjunction of features to be considered, exerting substantially greater demand on perceptual processes.

Participants were presented with 10 practice trials before undertaking the task, in which the correct answer and distinguishing features were highlighted to the participant after they had made their response. Test trials (*n*=60) were presented in a random order. All participants completed the task in a remote, ‘home’ environment via a subdomain on the lab website ((d01.eventmemory.org). Access to the task was restricted to desktop or laptop computers.

### 2.3 Statistical Analysis

Initially, data was screened for participants showing deviant task performance. No individuals met pre-registered criteria for exclusion, namely, being classed as an outlier (defined as more than 3 standard deviation (sd) from the population norm) in both accuracy (proportion) and mean response time (RT) in the low ambiguity condition. Five individuals (2 APOE33, 2 APOE34 and 1 APOE44) were classed as outliers in terms of accuracy. However, re-running analyses with these individuals removed did not qualitatively change outcomes from the study. Chi-squared tests were used to test group-differences in the distributions of gender, education, and family history of dementia.

Frequentist generalised-linear mixed effects models (GLMMs) were computed by maximum-pseudolikelihood estimation in R (Package: lme4, version1 -1.32). Equivalent Bayesian models were estimated using the ‘BRMS’ package in RStan (version 2.19.0). Frequentist statistics and Bayesian 95% credibility intervals (CIs) are reported for main effects and interaction terms, plus pairwise comparisons of interest (*p*-values adjusted using Holm-Bonferroni method for multiple comparisons). Bayesian CIs were computed by multiplying a contrast vector or matrix by all posterior samples returned by RStan. For contrasts with 1 degree of freedom, CIs were taken as the upper and lower limits that included the central 95% of contrasted samples. To produce a single interval for contrasts with more than 1 degree of freedom, contrasted posterior samples were multiplied by a rank-1 unit vector coincident with the contracted posterior mean before the upper and lower limits were calculated. Model parameter estimates and standard errors are uploaded to the project repository on the Open Science Framework, alongside anonymised trial-level datasets and scripts for running GLMMs in R (https://osf.io/qns2r/).

#### 2.3.1 Accuracy

The number of correct perceptual discriminations (outcome variable) was modelled in two GLLMs as a binomial process with a logit link function such that parameter estimates encode the probability of a correct response on each trial. For Bayesian model estimates, the prior for the intercept term was set to a logistic distribution with µ= 0 and s= 1. Priors for all other fixed effects were set to a Cauchy distribution with a location parameter (x_0_) of 0, and a scale parameter (y) of 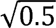^1^. Model 1a, testing for an effect of e4 status, included the following fixed effects: **1)** APOE e4 status (APOE33= 0, APOE e4+ = 1); **2)** Ambiguity (Low = 0, High = 1); **3)** z-standardised 5-year age band, - plus all possible interaction terms. Model 2a, testing for an effect of e4 gene dose, included the following fixed effects: **1)** APOE e4 gene dose modelled as a factor with two dummy coded predators (APOE34 vs APOE33, and APOE44 vs APOE33); **2)** Ambiguity (Low = 0, High = 1); **3)** z-standardised 5-year age band, - plus all possible interaction terms. Note, APOE genotype haplotype was modelled as a non-linear factor given that this structure yielded a lower AIC score (1373.0) when compared to a model that treated gene dose as a linear predictor (1374.1). Both Models 1a and 2a included random intercepts for each participant.

In addition to trial-level GLMMs, a standardised difference score (z (% correct high ambiguity trials - % correct low ambiguity trials)) was extracted to provide a single metric of conjunction-based perceptual discrimination after controlling for differences in basic perceptual ability, consistent with past literature (Gellersen et al., 2022). Genotype group differences were considered using a between-groups t-test (APOE33 vs. APOE e4+) and a one-way analysis of variance (ANOVA; APOE33 vs. APOE34 vs. APOE44) with age included as a covariate.

#### 2.3.2 Response time

Prior to analysis, RTs more than 3sd away from the individual’s personal mean RT were removed. Trial RT (in seconds) for correct decisions was subsequently modelled in two GLMMs with a log link function and the gamma distribution. For Bayesian models, the prior for the intercept term was set to a lognormal distribution with µ= 0 and o= 1. Priors for all other fixed effects were set to a Cauchy distribution with a location parameter (x_0_) of 0, and a scale parameter (y) of 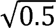. Model 1b included the same fixed- and random-effects predictors as Model 1a (for accuracy, see above). Similarly, Model 2b had the same fixed-and random-effects predictors as Model 2a with the exception that APOE haplotype was modelled by a single linear predictor indicating gene dose (APOE33=0, APOE34=1, APOE44=2). This was done as a single linear predictor yielded a better fit to the data (AIC=79124.6) compared with a model that treated APOE haplotype as a categorical variable (AIC=79129.2).

#### 2.3.3 Speed-accuracy trade-off

Exploratory analyses (not pre-registered) extracted the correlation between mean RT (correct responses) and mean proportion accuracy in the high ambiguity condition for each genotype group. This additional analysis was completed to examine if group differences in strategy for task completion (e.g., being slower to maintain accuracy) accounted for Genotype or Age effects.

## 3. Results

The distribution of ages is equivalent between APOE33 and APOE e4+ individuals (x^2^(2)=3.29, *p*=.350), however, there is a trend-level difference in the distribution of age-groups by APOE e4 gene dose (x^2^(3)=12.56, *p*=.051, with a higher proportion of the APOE44 group being in the 60 – 65 years age bracket. Genotype groups were also equivalent in gender, education level, and immediate (parent or sibling) family history of dementia (*p*>.05). Including gender in reported statistical models did not influence reported genotype effects (see Supplementary Materials for effects of Gender on task performance). Descriptive statistics for task performance (proportion accuracy, mean RT) are shown in table 2.

**Table 2.** Descriptive statistics of performance accuracy (%) and mean RT (correct trials only) on the Greebles “odd-one-out” task, shown by APOE e4 status and APOE e4 gene dose.

|  | **E4 status** |  | **E4 gene dose** |  |
| --- | --- | --- | --- | --- |
| **Accuracy (%)** | **APOEE33** | **APOE e4+** | **APOE34** | **APOE44** |
| Low Ambiguity | 98.83 ± 2.87 | 99.20 ± 2.23 | 99.28±1.94 | 98.50 ± 3.01 |
| High Ambiguity | 95.74 ± 6.67 | 95.52 ± 6.80 | 96.28 ± 5.70 | 92.97 ± 9.46 |
| **Response time (s)** |  |  |  |  |
| Low Ambiguity | 4.75 ± 2.61 | 5.17 ± 3.99 | 5.24 ± 4.23 | 4.92 ± 2.90 |
| High Ambiguity | 6.46 ± 4.33 | 7.12 ± 5.63 | 7.18 ± 5.88 | 6.90 ± 4.54 |

### 3.1 Effect of e4 status on response accuracy

Model 1a (626 observations) reports a significant main effect of increasing trial ambiguity on accuracy, *F*(1, 617)= 140.90, *p*<.001; Bayesian 95% CI: [-3.77, -2.72]. As expected, the probability of a correct response is higher for low relative to high ambiguity trials. In addition, there is a significant main effect of Age, *F*(1, 617)= 10.86, *p*=.001; Bayesian 95% CI: [-1.89, -.28]. The main effect of e4 status, *F*(1, 617)= .03, *p=.*865; Bayesian 95% CI: [-.49, .98], however, was non-significant, as were all interaction terms in the model (*p* >.05; Bayesian 95% CIs cross 0). When considering *z-*standardised difference scores, there was no significant difference between APOE33 (M=-3.09 ± 5.37) and APOE e4+ (*M=-*3.68 ± 5.73); *F*(1, 305)=.87, *p*=.352, *η^2^_ρ_* =.00.

### 3.2 Effect of e4 gene dose on response accuracy

Model 2a (626 observations) again reports significant main effects of trial Ambiguity, *F*(1, 613) = 138.03, p<.001, Bayesian 95% CI: [-5.84, -.4.04] and Age, *F*(1, 613)=11.11, *p*<.001, Bayesian 95% CI: [-4.56, -.67]. The main effect of APOE gene dose is also significant, *F*(2,613) = 3.75, *p*=.025, Bayesian 95% CI: [.07, 1.90]. Planned contrasts (holm-Bonferroni adjusted; Bayesian), however, are not indicative of a significant pairwise difference between APOE33 and APOE34 (*p*=.436, critical a=.016, Bayesian 95% CI: [-.48, .99]), APOE33 and APOE44 (*p*=.096, critical a=.013, Bayesian 95% CI: [-2.12, .24]), or APOE34 and APOE44 groups (*p*=.035, critical a=.010, Bayesian 95% CI: [-2.39, .03]). The Ambiguity x APOE gene dose interaction (Figure 2) was non-significant (*p*=.258, Bayesian 95% CI: [-.18, 1.02], as were all other interaction terms in the model (*p*>.05, Bayesian 95% CI include 0). The Ambiguity x Age x APOE gene dose interaction (*p*=.199, Bayesian 95% CI: [-.26, 1.20]) is shown in Supplementary Figure 1.

**Figure 1.**
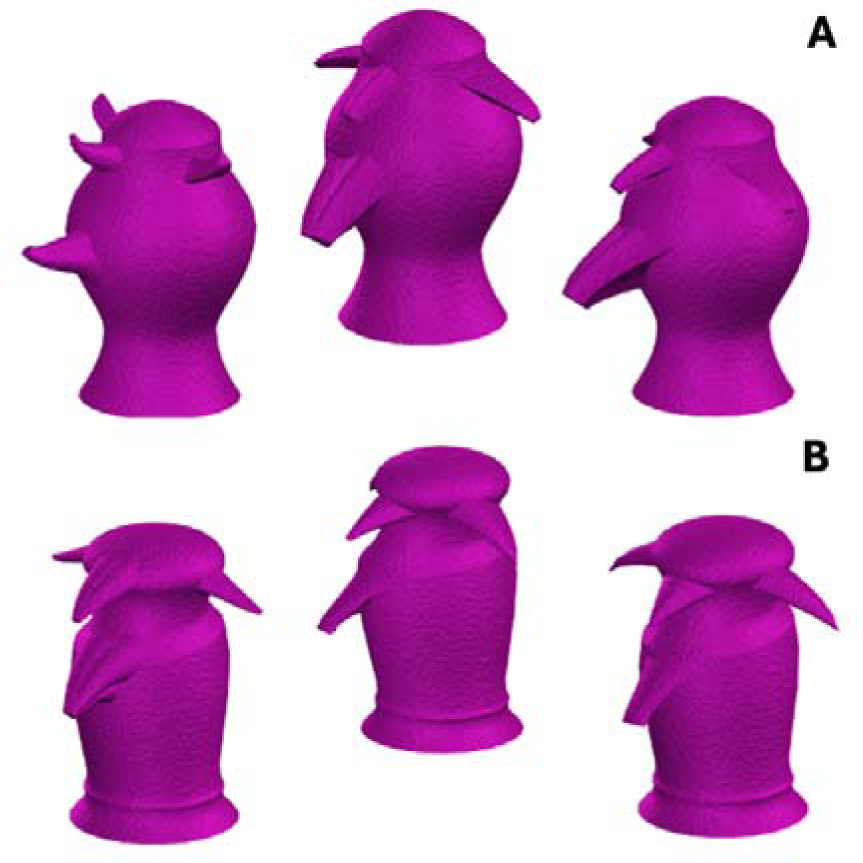
Example stimuli arrays in the Greebles ‘odd-one-out’ task. Greebles in the low ambiguity condition (A) share a small proportion of perceptual features, relative to Greebles in the high ambiguity condition (B). [Colour print]

**Figure 2.**
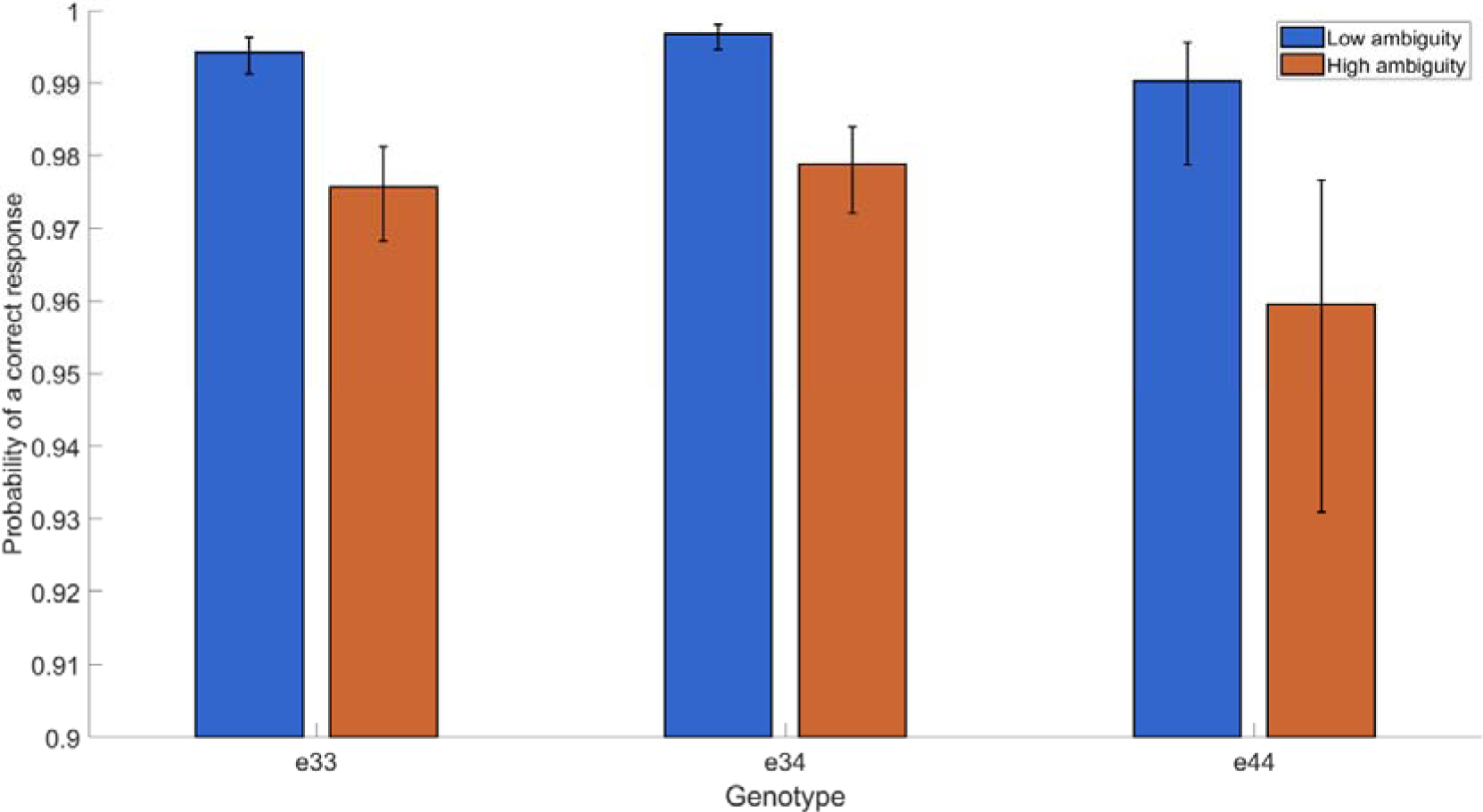
Frequentist model estimates of probability of correct response on low and high ambiguity trials on the Greebles ‘odd-one-out’ task, separated by APOE e4 gene dose. [Colour print]

Of note, GLMM 2a reports a significant main effect of gene dose on task accuracy (*p*=.025, Bayesian 95% CI: [.07, 1.90]) despite ceiling effects in the low ambiguity condition. Critically, these ceiling effects may also skew model estimates of interaction effects. However, whilst the Dose x Ambiguity interaction was non-significant in GLMM 2a (*p*=.258, Bayesian 95% CI: [-.18, 1.02]), there was a significant effect of APOE e4 gene dose on Z-standardised difference scores which reflect the cost of increasing trial ambiguity on task accuracy, *F*(2, 301)=3.88, *p*=.022, *η^2^_p_*=.03. Following Holm-Bonferroni adjustment, post-hoc comparisons suggested there was a significant difference in Z-standardised difference scores between e33 carriers (*M=-*3.09 ± 5.37) and e44 carriers (*M=-*5.83 ± 8.47; *p*=.025) and e34 carriers and e44 carriers (*M=-*3.12 ± 4.62; *p*=.025). The main effect of Age and Age x APOE haplotype interaction were both non-significant (*p*>.05) on z-standardised difference scores. Note, analyses of Z-standardised difference scores were pre-registered in advance and replicate the methods of a past paper utilising this paradigm [9].

### 3.3 Effect of e4 status on response time

Model 1b (17962 observations) reports a significant main effect of trial ambiguity on response time, *F*(1, 17952) = 1895.95, *p*<.001; Bayesian 95% CI: [.58, .63]. As expected, RTs are slower in the high ambiguity relative to the low ambiguity condition (table 2). There is also a significant main effect of Age, *F*(1, 17952)=112.89, *p*<.001, Bayesian 95% CI: [.36, .65], with RTs slowing with increasing age. However, the main effect of APOE e4 status is non-significant, *F*(1, 17952)=1.53, *p*=2.16, Bayesian 95% CI: [-.10, .22]. There was a significant Age x APOE e4 status interaction, *F*(1,17952)=12.95, *p*<.001; Bayesian 95% CI: [.01, .33]. As can be seen in Figure 3, in the younger age-group, the APOE e4+ responded faster than the APOE e33 control group across both low and high ambiguity trials, whereas this pattern was reversed in adults aged 60 years and older. The Ambiguity x Age interaction was also significant, *F*(1,17952)=8.79, *p*=.003; Bayesian 95% CI: [.02, .07], with age-related slowing greater on high ambiguity relative to low ambiguity trials. Both the Ambiguity x APOE e4 status and 3-way Ambiguity x Age x APOE e4 status interactions, however, were non-significant (*p*>.05, Bayesian 95% CI include 0].

**Figure 3.**
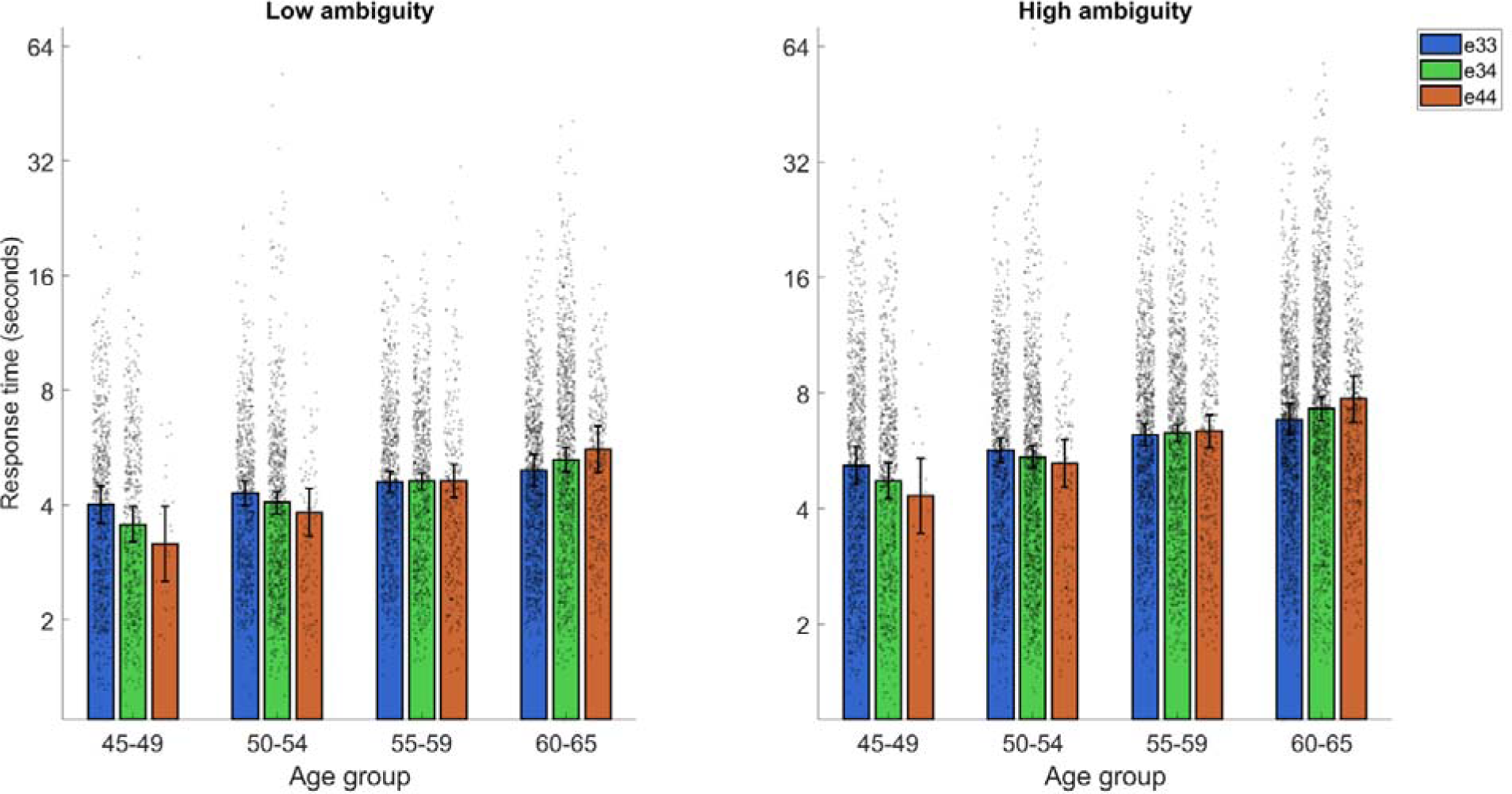
Frequentist model estimates of response time (RT) for correct perceptual judgements on low and high ambiguity trials, separated by APOE e4 gene dose and 5-year age-band. [Colour print]

### 3.4 Effect of e4 gene dose on response time

As in Model 1b, Model 2b (17962 observations) reports a significant main effects of trial Ambiguity on RT, *F*(1, 17952) = 1895.46, *p*<.001; Bayesian 95% CI: [.58, .63] and Age, *F*(1, 17952)=112.32, *p*<.001; Bayesian 95% CI: [.32, .64]. The main effect of Gene Dose, however, is non-significant, *F*(1, 17952)=.09, *p*=.758; Bayesian 95% CI: [-.13, 11]. There was a significant Age by Ambiguity interaction, *F*(1, 17952)= 8.79, *p*=.003; Bayesian 95% CI: [.01, .07] and Age x APO*E* e4 gene dose interaction, *F*(1,17952)=9.75, *p*=.002; Bayesian 95% CI: [.00, .25] (Figure 3). Genotype effects on RT reversed with increasing age, and this effect was stronger in APOE e4 homozygous group relative to heterozygous individuals (see Figure 3). All other interaction terms were non-significant (*p*>.05; Bayesian CIs cross 0).

### 3.5 Speed-accuracy trade-off

There was a small, non-significant correlation between proportion accuracy and mean RT to correct trials in all three genotype groups: APOE33 - ρ =-.16, *p=.*052; APOE34 - ρ =.08, *p=.*354; APOE44 - ρ =.04, *p=.*827. This suggests there were no differential speed/accuracy strategies adopted between genotypes.

## 4. Discussion

The present study aimed to establish whether possession of an APOE e4 genetic risk variant for dementia is associated with impaired perceptual discrimination in mid-life. In addition, the number of APOE44 individuals present in this online study facilitated novel exploration of gene-dose effects. Methods employed a remote Greebles ‘odd-one-out’ task, with performance on this task correlated with structure and function of the MTL [11] - a brain-region compromised in early AD.

Current results show only APOE44 individuals are impaired in their perceptual discrimination abilities by mid-life, with this highest-risk group being less accurate in their judgements than APOE34 individuals and an APOE33 control group. Furthermore, whilst the Ambiguity by Genotype interaction was not significant, standardised difference scores between accuracy on high and low ambiguity trials indicate APOE44 individuals show greater cost of increasing perceptual overlap. Cross-sectional analyses revealed slopes of age-related slowing of reaction times increased linearly with the number of APOE e4 risk alleles carried, across both low and high ambiguity trials. This may indicate that whilst APOE34 carriers show intact cognitive performance into the 6^th^ decade, reduced efficiency of perceptual judgements was recorded in the oldest age-band (60-65 years) and may reflect subtle disadvantages emerging with increasing age.

Observed performance disadvantages in mid-age APOE44 are consistent with prior studies reporting perceptual discrimination tasks are sensitive to heightened familial risk of AD in mid-life [26], plus distinguish older adults at risk of converting to dementia based on neuropsychological test scores [9,10]. This supports the utility of paradigms which tax high-level perceptual processing as early cognitive markers of AD risk. Current results are not inconsistent with past research evidencing equivalent perceptual discrimination in young heterozygous APOE e4 carriers [25] as accuracy was maintained in our APOE34 group, coupled with quicker RTs until age 60. This previous neuroimaging study [25], however, did not include APOE44 individuals, nor examine speed of perceptual decision-making as an outcome of interest. Furthermore, reported cognitive effects of APOE e4 are not uniform across the lifespan [20,34], with some studies reporting APOE e4 advantages in MTL-dependent [35:36] and speed-dependent tasks [37;38] in younger years. Given the pattern of age-related change in perceptual processing speed observed here, younger APOE34 and APOE44 may be expected to show stepwise advantages in high-speed decision-making in early adulthood.

This study provides the first evidence that APOE e4 deficits in perceptual discrimination emerge as a function of the number of risk alleles carried in mid-life, with disadvantages primarily seen for homozygote individuals. Due to the relatively low (∼2%) prevalence of APOE44 individuals in the population [39], few lab-based studies have considered the impact of APOE e4 zygosity. Paradigms which place significant challenge on MTL-function, however, support the presence of a gene-dose effect from the 5^th^ decade. Specifically, a linear disadvantage effect of APOE e4 was observed in cognitively healthy adults aged 40-60 years old when recall performance was stressed by a 7-day delay between learning and test (as opposed to 30-minutes) [40]. In the same sample, a detrimental effect of APOE e4 zygosity was also reported in the ability to harness audio memory to guide attention [41]. Standardised neuropsychological assessment is more commonly employed by large-scale birth cohorts. Examination of longitudinal trends suggest only APOE44 individuals show a significant decline in episodic memory from age 43-69 years (relative to APOE33 counterparts [42], although a comparable, non-significant trend was seen when looking at APOE34 carriers. This effect was not seen on tests of processing speed, suggesting disadvantageous effects of APOE4 gene dose may be heightened in tasks probing tau-sensitive MTL-regions.

The PRc is one brain region involved in our ability to process objects as a conjunction of perceptual features [8; 43], for example the high-ambiguity trials in this Greebles ‘odd-one-out’ task. APOE44 individuals showed increased cost of higher trial ambiguity, which may suggest altered PRc function in this highest-risk group. Given that the PRc is the site of earliest neurofibrillary tau deposition, reported APOE e4 genotype differences may indicate homozygote APOE e4 carriers are showing premature, toxic-gain of tau neuropathology [22:24]. Recent evidence collating post-mortem neuropathology, clinical, and biomarker data from 5 cohorts suggests the predictability of biomarker trajectories and symptom onset, plus near-complete penetrance of biomarker positivity in homozygous individuals is comparable to autosomal forms of AD [30]. Indeed, models of biomarker trajectories suggest that tau positivity begins to increase substantially in APOE44 individuals around age 55, in agreement with theoretical accounts of the current results. As such, perceptual discrimination errors may reflect that even by mid-adulthood, APOE44 represent an AD-prodrome. Perceptual disadvantages in APOE44 carriers may also reflect early functional reorganisation of MTL-network, with increasing age correlating with poorer mnemonic discrimination at short memory delays and reduced specialisation of PRc and alERc for processing objects relative to scenes [13].

APOE44 carriers, and to a lesser extent APOE34 carriers showed increased age-related slowing in perceptual judgments. Curiously, APOE e4 carriers as a collective appeared faster in early mid-life. This may confer support for the antagonistic pleiotropy account of APOE; that carriers may show advantages in tasks requiring sustained attention in younger years [37:38, 44]. This result was consistent across low and high-ambiguity trials suggesting efficiency may reflect a domain-general process like processing speed, rather than exclusively representing the integrity of the PRc. In older adults, processing speed was reported to decline as a function of APOE e4 zygosity [45], however, gene-dose effects in this domain were not reported in mid-age [42]. There has been limited consideration of reaction time in perceptual judgement tasks; however, Jiang and colleagues [10] reported no difference in discrimination times between cognitively healthy older adults and those with impairments in global cognition. In the current study there was no suggestion that genotype groups were preferentially prioritising accuracy at the expense of RT or vice versa. However, as the Greebles perceptual discrimination task is sensitive to age-effects by mid-life[46], it may be that APOE e4 carriers as a collective are more vulnerable to accelerated ageing.

Of note, accuracy in this study was close to ceiling for both low and high ambiguity trials which may have limited our sensitivity to genotype effects. Stimuli are central to perceptual discrimination tasks; ‘Greeble’ objects are relatively humanoid in their appearance which may benefit our ability to distinguish between them. A recent study manipulated the spatial arrangement and type of feature extracted from letter stimuli to create a more abstract, 6-item ‘odd-one-out’ task [8]. Accuracy on this task distinguished healthy older adults from individuals diagnosed with mild AD and correlated with volume of the PRc and entorhinal cortex. Furthermore, the extent to which viewpoint differs when distinguishing between stimuli is correlated with increased blood-oxygen-level-dependent (BOLD) response in the PRc [11], suggesting mental rotation may contribute to differences in task performance. Future research may consider whether the sensitivity of perceptual judgement tasks could be elevated by further challenging detailed perceptual processing and the role of mental rotation in task performance.

It must be acknowledged that whilst recruiting through NIHR BioResource allowed us access to a greater number of APOE44 individuals than prior experimental studies, the study remained underpowered to reliably detect small (Cohen’s f = .15) differences between APOE e4 genotypes . In addition, whilst genotype groups were gender-matched with a range of educational backgrounds, our sample was not ethnically diverse. Embedding paradigms hypothesised to be sensitive to ‘preclinical’ neurodegenerative disease, such as MTL-dependent perceptual discrimination tasks, in biomarker rich, longitudinal population cohorts will further facilitate our understanding of how risk manifests across the lifespan. Digital, scalable, and remote assessments can accelerate this work. Furthermore, such studies can further our understanding of how cognitive assessments can supplement prediction of very early, sub-threshold biomarker development [2].

To conclude, perceptual discrimination tasks appear sensitive to the deleterious effects of APOE44 by mid-life. This potentially reflects increased vulnerability of this group to premature tau neuropathology, with recent evidence reporting nearly all APOE44 were biomarker positive by age 55 [30]. Mid-adulthood is a time of strategic importance for slowing the development of neurodegenerative conditions. Future research can harness measures of perceptual discrimination to facilitate cost-effective screening for clinical trials targeting the preclinical stages of disease, plus promote individualised interventions to mitigate risk prevention. First, further evidence elucidating the relationship between perceptual discrimination, tau neuropathology, and APOE in mid-life is needed.

## Data Availability

Model parameter estimates and standard errors are uploaded to the project repository on the Open Science Framework, alongside anonymised trial-level datasets and scripts for running statistical models in R (https://osf.io/qns2r/).

## 5. Acknowledgments

We thank NIHR BioResource volunteers for their participation, and gratefully acknowledge NIHR BioResource centres, NHS Trusts, and staff for their contribution. We thank the National Institute for Health and Care Research, NHS Blood and Transplant, and Health Data Research UK as part of the Digital Innovation Hub Programme. The views expressed are those of the author(s) and not necessarily those of the NHS, the NIHR or the Department of Health and Social Care.

## Footnotes

1 Prior 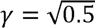 is in keeping with expectation of medium-sized effects.

## 8. References

1. Bastin, C., & Delhaye, E. (2023). Targeting the function of the transentorhinal cortex to identify early cognitive markers of Alzheimer’s disease. *Cognitive, Affective*, & Behavioral Neuroscience, 23(4), 986–996. 10.3758/s13415-023-01093-5

2. Elman, J. A., Panizzon, M. S., Gustavson, D. E., Franz, C. E., Sanderson-Cimino, M. E., Lyons, M. J., & Kremen, W. S. (2020). Amyloid-β positivity predicts cognitive decline but cognition predicts progression to amyloid-β positivity. Biological Psychiatry, 87(9), 819– 828. 10.1016/j.biopsych.2019.12.021

3. Thomas, K. R., Bangen, K. J., Weigand, A. J., Edmonds, E. C., Wong, C. G., Cooper, S., … & Alzheimer’s Disease Neuroimaging Initiative. (2020). Objective subtle cognitive difficulties predict future amyloid accumulation and neurodegeneration. Neurology, 94(4), e397-e406. doi: 10.1212/WNL.0000000000008838.

4. Lancaster, C., Tabet, N., & Rusted, J. (2017). The elusive nature of APOE ε4 in mid-adulthood: Understanding the cognitive profile. Journal of the International Neuropsychological Society, 23(3), 239–253. 10.1017/S1355617716000990

5. Mortamais, M., Ash, J. A., Harrison, J., Kaye, J., Kramer, J., Randolph, C., … & Ritchie, K. (2017). Detecting cognitive changes in preclinical Alzheimer’s disease: A review of its feasibility. Alzheimer’s & dementia, 13(4), 468–492. 10.1016/j.jalz.2016.06.2365

6. Jutten, R. J., Sikkes, S. A., Amariglio, R. E., Buckley, R. F., Properzi, M. J., Marshall, G. A., … & Alzheimer Disease Neuroimaging Initiative. (2021). Identifying sensitive measures of cognitive decline at different clinical stages of Alzheimer’s disease. Journal of the International Neuropsychological Society, 27(5), 426-438. 10.1017/S1355617720000934

7. Gaynor, L. S., Cid, R. E. C., Penate, A., Rosselli, M., Burke, S. N., Wicklund, M., … & Bauer, R. M. (2019). Visual object discrimination impairment as an early predictor of mild cognitive impairment and Alzheimer’s disease. Journal of the International Neuropsychological Society, 25(7), 688–698. 10.1017/S1355617719000316

8. Frei, M., Berres, M., Kivisaari, S. L., Henzen, N. A., Monsch, A. U., Reinhardt, J., Blatow, M., Kressig, R. W., & Krumm, S. (2023). Can you find it? Novel oddity detection task for the early detection of Alzheimer’s disease. Neuropsychology, 37(7), 717–740. 10.1037/neu0000859

9. Gellersen, H. M., Trelle, A. N., Farrar, B. G., Coughlan, G., Korkki, S. M., Henson, R. N., & Simons, J. S. (2023). Medial temporal lobe structure, mnemonic and perceptual discrimination in healthy older adults and those at risk for mild cognitive impairment. Neurobiology of Aging, 122, 88–106. 10.1016/j.neurobiolaging.2022.11.004

10. Jiang, L., Robin, J., Shing, N., Mazloum-Farzaghi, N., Ladyka-Wojcik, N., Balakumar, N., … & Olsen, R. K. (2024). Impaired perceptual discrimination of complex objects in older adults at risk for dementia. Hippocampus, 34(4), 197–203. 10.1002/hipo.23598

11. Barense, M. D., Henson, R. N., Lee, A. C., & Graham, K. S. (2010). Medial temporal lobe activity during complex discrimination of faces, objects, and scenes: Effects of viewpoint. Hippocampus, 20(3), 389–401. 10.1002/hipo.20641

12. Lawrence, A. V., Cardoza, J., & Ryan, L. (2020). Medial temporal lobe regions mediate complex visual discriminations for both objects and scenes: A process[based view. Hippocampus, 30(8), 879–891. 10.1002/hipo.23203

13. Berron, D., Neumann, K., Maass, A., Schütze, H., Fliessbach, K., Kiven, V., … & Düzel, E. (2018). Age-related functional changes in domain-specific medial temporal lobe pathways. Neurobiology of aging, 65, 86–97. 10.1016/j.neurobiolaging.2017.12.030

14. Ferko, K. M., Blumenthal, A., Martin, C. B., Proklova, D., Minos, A. N., Saksida, L. M., … & Köhler, S. (2022). Activity in perirhinal and entorhinal cortex predicts perceived visual similarities among category exemplars with highest precision. elife, 11, e66884. 10.7554/eLife.66884

15. Bussey, T. J., Saksida, L. M., & Murray, E. A. (2005). The perceptual-mnemonic/feature conjunction model of perirhinal cortex function. The Quarterly Journal of Experimental Psychology Section B, 58(3-4b), 269-282. 10.1080/02724990544000004

16. Martin, C. B., & Barense, M. D. (2023). Perception and memory in the ventral visual stream and medial temporal lobe. Annual review of vision science, 9(1), 409–434. 10.1146/annurev-vision-120222-014200

17. Braak, H., & Braak, E. (1991). Neuropathological stageing of Alzheimer-related changes. Acta neuropathologica, 82(4), 239–259. 10.1007/BF00308809

18. Hirni, D. I., Kivisaari, S. L., Monsch, A. U., & Taylor, K. I. (2013). Distinct neuroanatomical bases of episodic and semantic memory performance in Alzheimer’s disease. Neuropsychologia, 51(5), 930–937. 10.1016/j.neuropsychologia.2013.01.013

19. Farrer, L. A., Cupples, L. A., Haines, J. L., Hyman, B., Kukull, W. A., Mayeux, R., … & Van Duijn, C. M. (1997). Effects of age, sex, and ethnicity on the association between apolipoprotein E genotype and Alzheimer disease: a meta-analysis. Jama, 278(16), 1349–1356. doi:10.1001/jama.1997.03550160069041

20. O’Donoghue, M. C., Murphy, S. E., Zamboni, G., Nobre, A. C., & Mackay, C. E. (2018). APOE genotype and cognition in healthy individuals at risk of Alzheimer’s disease: A review. cortex, 104, 103–123. 10.1016/j.cortex.2018.03.025

21. Steele, O. G., Stuart, A. C., Minkley, L., Shaw, K., Bonnar, O., Anderle, S., … & King, S. (2022). A multi[hit hypothesis for an APOE4[dependent pathophysiological state. European Journal of Neuroscience, 56(9), 5476–5515. 10.1111/ejn.15685

22. Daly, J., De Luca, F., Berens, S. C., Field, A. P., Rusted, J. M., & Bird, C. M. (2024). The effect of apolipoprotein E genotype on spatial processing in humans: a meta-analysis and systematic review. Cortex. 10.1016/j.cortex.2024.05.006

23. Young, C. B., Johns, E., Kennedy, G., Belloy, M. E., Insel, P. S., Greicius, M. D., … & A4 Study Team. (2023). APOE effects on regional tau in preclinical Alzheimer’s disease. Molecular neurodegeneration, 18(1), 1. 10.1186/s13024-022-00590-4

24. Steward, A., Biel, D., Dewenter, A., Roemer, S., Wagner, F., Dehsarvi, A., … & Franzmeier, N. (2023). ApoE4 and connectivity-mediated spreading of tau pathology at lower amyloid levels. JAMA neurology, 80(12), 1295–1306. doi:10.1001/jamaneurol.2023.4038

25. Therriault, J., Benedet, A. L., Pascoal, T. A., Mathotaarachchi, S., Chamoun, M., Savard, M., … & Rosa-Neto, P. (2020). Association of apolipoprotein E ε4 with medial temporal tau independent of amyloid-β. JAMA neurology, 77(4), 470–479. doi:10.1001/jamaneurol.2019.4421

26. Shine, J. P., Hodgetts, C. J., Postans, M., Lawrence, A. D., & Graham, K. S. (2015). APOE-ε4 selectively modulates posteromedial cortex activity during scene perception and short-term memory in young healthy adults. Scientific Reports, 5(1), 16322. 10.1038/srep16322

27. Mason, E. J., Hussey, E. P., Molitor, R. J., Ko, P. C., Donahue, M. J., & Ally, B. A. (2017). Family history of Alzheimer’s disease is associated with impaired perceptual discrimination of novel objects. Journal of Alzheimer’s Disease, 57(3), 735–745. doi: 10.3233/JAD-160772

28. Barense, M. D., Gaffan, D., & Graham, K. S. (2007). The human medial temporal lobe processes online representations of complex objects. Neuropsychologia, 45(13), 2963–2974. 10.1016/j.neuropsychologia.2007.05.023

29. Bertram L, McQueen MB, Mullin K, Blacker D, Tanzi RE. (2007) “Systematic meta-analyses of Alzheimer disease genetic association studies: the AlzGene database.” Nat Genet 39(1): 17–23. 10.1038/ng1934.

30. Genin, E., Hannequin, D., Wallon, D., Sleegers, K., Hiltunen, M., Combarros, O., … & Campion, D. (2011). APOE and Alzheimer disease: a major gene with semi-dominant inheritance. Molecular psychiatry, 16(9), 903–907. 10.1038/mp.2011.52

31. Fortea, J., Pegueroles, J., Alcolea, D., Belbin, O., Dols-Icardo, O., Vaqué-Alcázar, L., … & Montal, V. (2024). APOE4 homozygozity represents a distinct genetic form of Alzheimer’s disease. Nature medicine, 1-8. 10.1038/s41591-024-02931-w

32. Suri, S., Heise, V., Trachtenberg, A. J., & Mackay, C. E. (2013). The forgotten APOE allele: a review of the evidence and suggested mechanisms for the protective effect of APOE [2. Neuroscience & Biobehavioral Reviews, 37(10), 2878–2886. 10.1016/j.neubiorev.2013.10.010

33. Kim, H., Devanand, D. P., Carlson, S., & Goldberg, T. E. (2022). Apolipoprotein E genotype e2: neuroprotection and its limits. Frontiers in Aging Neuroscience, 14, 919712. 10.3389/fnagi.2022.919712

34. Wisdom, N. M., Callahan, J. L., & Hawkins, K. A. (2011). The effects of apolipoprotein E on non-impaired cognitive functioning: a meta-analysis. Neurobiology of aging, 32(1), 63–74. 10.1016/j.neurobiolaging.2009.02.003

35. Stening, E., Persson, J., Eriksson, E., Wahlund, L. O., Zetterberg, H., & Söderlund, H. (2016). Apolipoprotein E [4 is positively related to spatial performance but unrelated to hippocampal volume in healthy young adults. Behavioural Brain Research, 299, 11–18. 10.1016/j.bbr.2015.11.006

36. Mondadori, C. R., de Quervain, D. J. F., Buchmann, A., Mustovic, H., Wollmer, M. A., Schmidt, C. F., … & Henke, K. (2007). Better memory and neural efficiency in young apolipoprotein E ε4 carriers. Cerebral cortex, 17(8), 1934–1947. 10.1093/cercor/bhl103

37. Marchant, N. L., King, S. L., Tabet, N., & Rusted, J. M. (2010). Positive effects of cholinergic stimulation favor young APOE [4 carriers. Neuropsychopharmacology, 35(5), 1090–1096. 10.1038/npp.2009.214

38. Rusted, J. M., Evans, S. L., King, S. L., Dowell, N., Tabet, N., & Tofts, P. S. (2013). APOE e4 polymorphism in young adults is associated with improved attention and indexed by distinct neural signatures. Neuroimage, 65, 364–373. 10.1016/j.neuroimage.2012.10.010

39. Belloy, M. E., Napolioni, V., & Greicius, M. D. (2019). A quarter century of APOE and Alzheimer’s disease: progress to date and the path forward. Neuron, 101(5), 820–838. 10.1016/j.neuron.2019.01.056

40. Zimmermann, J. F., & Butler, C. R. (2018). Accelerated long-term forgetting in asymptomatic APOE ε4 carriers. The Lancet Neurology, 17(5), 394–395. 10.1016/S1474-4422(18)30078-4

41. Zimmermann, J., Alain, C., & Butler, C. (2019). Impaired memory-guided attention in asymptomatic APOE4 carriers. Scientific Reports, 9(1), 8138. 10.1038/s41598-019-44471-1

42. Rawle, M. J., Davis, D., Bendayan, R., Wong, A., Kuh, D., & Richards, M. (2018). Apolipoprotein-E (Apoe) ε4 and cognitive decline over the adult life course. Translational psychiatry, 8(1), 18. 10.1038/s41398-017-0064-8

43. Delhaye, E., Bahri, M. A., Salmon, E., & Bastin, C. (2019). Impaired perceptual integration and memory for unitized representations are associated with perirhinal cortex atrophy in Alzheimer’s disease. Neurobiology of Aging, 73, 135–144. 10.1016/j.neurobiolaging.2018.09.021

44. Atkinson, R. Á., Gaysina, D., & Rusted, J. M. (2019). Responses to executive demand in young adulthood differ by APOE genotype. Behavioural Brain Research, 360, 158–168. 10.1016/j.bbr.2018.11.033

45. Marioni, R. E., Campbell, A., Scotland, G., Hayward, C., Porteous, D. J., & Deary, I. J. (2016). Differential effects of the APOE e4 allele on different domains of cognitive ability across the life-course. European Journal of Human Genetics, 24(6), 919–923. 10.1038/ejhg.2015.210

46. Gellersen, H. M., McMaster, J., Abdurahman, A., & Simons, J. S. (2024). Demands on perceptual and mnemonic fidelity are a key determinant of age-related cognitive decline throughout the lifespan. Journal of Experimental Psychology: General, 153(1), 200. doi: 10.1037/xge0001476

