## Supplementary Materials for "Perceptual discrimination of complex objects: APOE e4 gene-dose effects in mid-life"

**Supplementary Figure 1.** *Frequentist model estimates of probability of correct response on low and high ambiguity trials on the Greebles ‘odd-one-out’ task, separated by APOE e4 gene dose and 5-year age-band.*

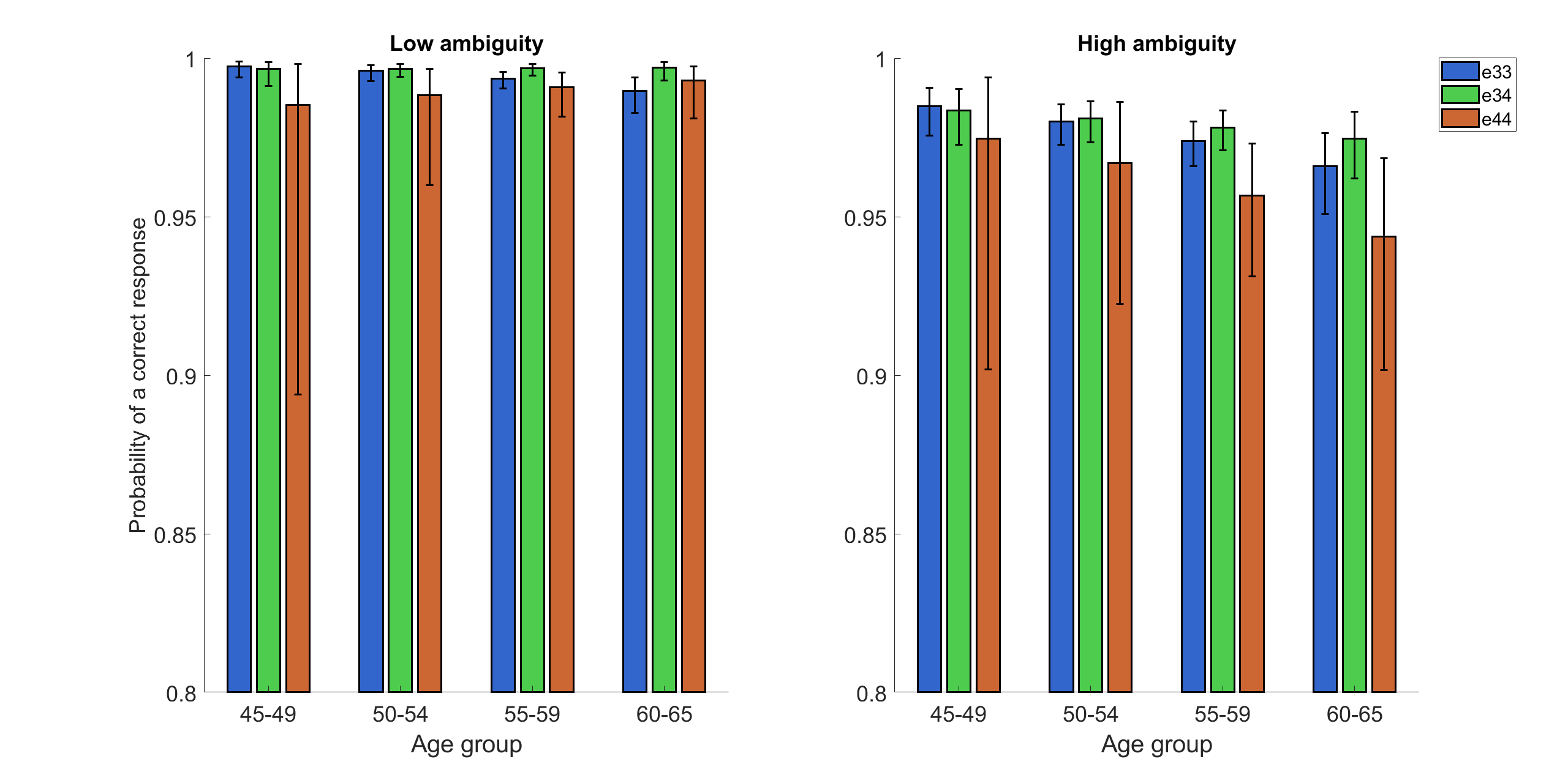

**Supplementary Table 1.** Descriptive statistics of performance accuracy (%) and mean RT (correct trials only) on the Greebles “odd-one-out” task shown by Age

|  | **5-year Age Group (years)** | | | |
| --- | --- | --- | --- | --- |
| **Accuracy (%)** | **45-49** | **50-54** | **55-59** | **60-65** |
| Low Ambiguity | 99.71 ± 1.07 | 98.94 ± 2.31 | 98.89 ± 2.47 | 98.82 ± 3.19 |
| High Ambiguity | 97.81 ± 3.81 | 95.74 ± 5.92 | 95.18 ± 7.90 | 94.68 ± 7.32 |
| **Response time (s)** |  |  |  |  |
| Low Ambiguity | 4.12 ± 2.41 | 4.34 ± 2.89 | 4.92 ± 2.77 | 5.56 ± 3.31 |
| High Ambiguity | 5.52 ± 3.70 | 5.67 ± 4.12 | 6.71 ± 4.10 | 7.76 ± 5.20 |

**Supplementary Results**

**1. Inclusion of Gender as main and interactive effect in model of response accuracy.**

Gender (0=male, 1=female) was included as an additional predictor in model 1a. The main effect of Gender, *F*(1, 609)= .92, *p=.*338; Bayesian 95% CI: [-.25, 1] was non-significant. Frequentist estimates support a significant Ambiguity x Gender interaction, however, this was not reflected in Bayesian 95% CI, *F*(1, 609)= 4.02, *p=.*045; Bayesian 95% CI: [-.90, .25] (shown in Supplementary Figure 2). All other interaction terms in the model including Gender were non-significant (*p* >.05; Bayesian 95% CIs cross 0).

**Supplementary Figure 2.** *Frequentist model estimates of probability of correct response on low and high ambiguity trials on the Greebles ‘odd-one-out’ task, separated by gender.*

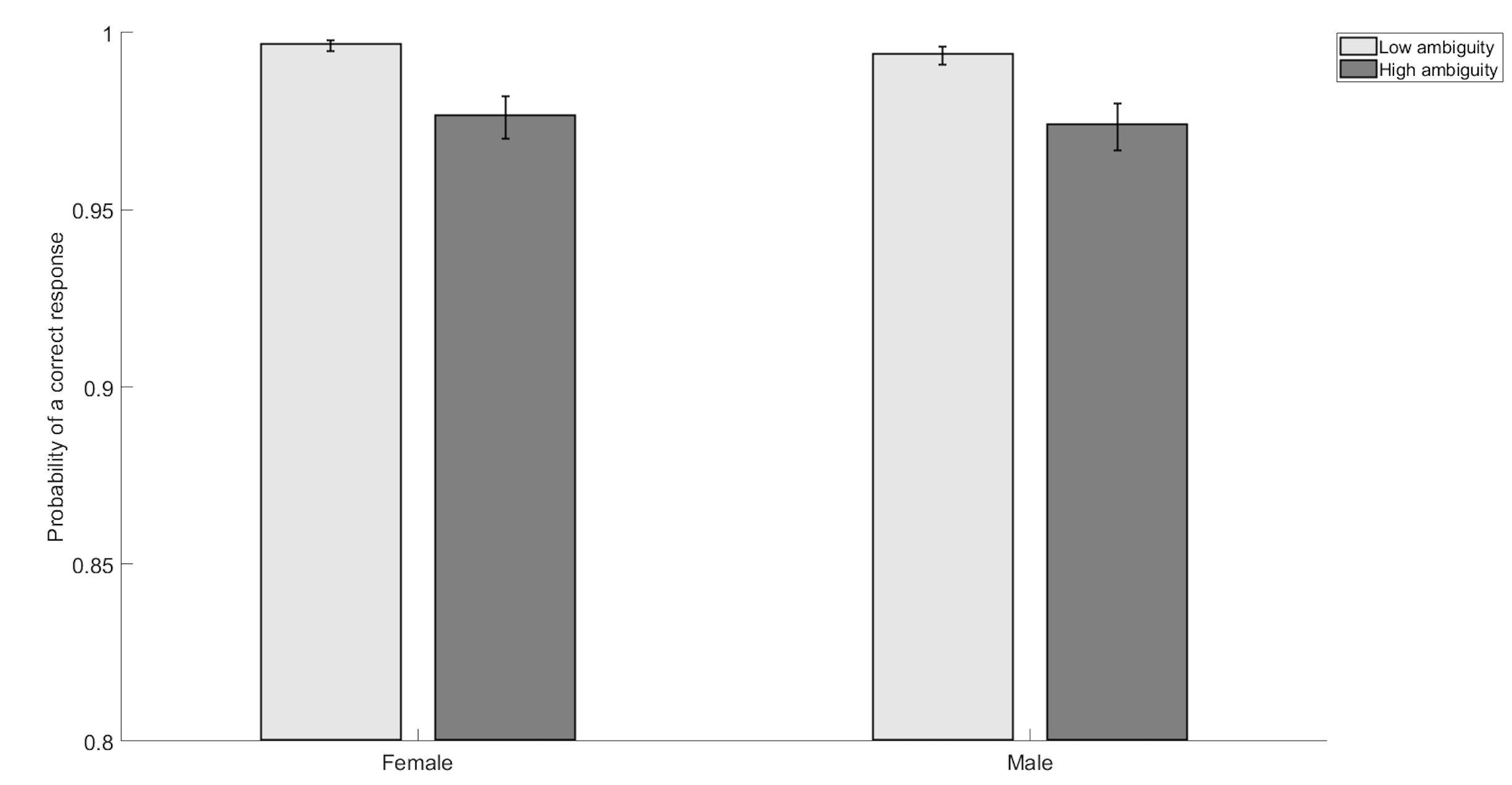

**2. Inclusion of Gender as main and interactive effect in model of response speed.**

Gender (0=male, 1=female) was included as an additional predictor in model 1b. The main effect of Gender, *F*(1, 17944)=.03, *p*=.862, Bayesian 95% CI: [-.07, .16] was non-significant. Frequentist statistics report significant Gender x Age interaction, *F*(1,17944)=10.47, *p*=.001; Bayesian 95% CI: [-.03, .20] and Gender x Age x Ambiguity interaction, *F*(1,17944)=3.95, *p*=.047; Bayesian 95% CI: [-.02, .06] but note Bayesian 95% CIs cross 0. Supplementary Figure 3 shows this 3-way interaction, suggesting females show a greater slope of age-related slowing, particularly in the high ambiguity condition. All other interaction terms in the model including Gender were non-significant (*p* >.05; Bayesian 95% CIs cross 0).

**Supplementary Figure 3.** *Frequentist model estimates of RT for correct trials on low and high ambiguity trials on the Greebles ‘odd-one-out’ task, separated by gender and 5-year age-band.*

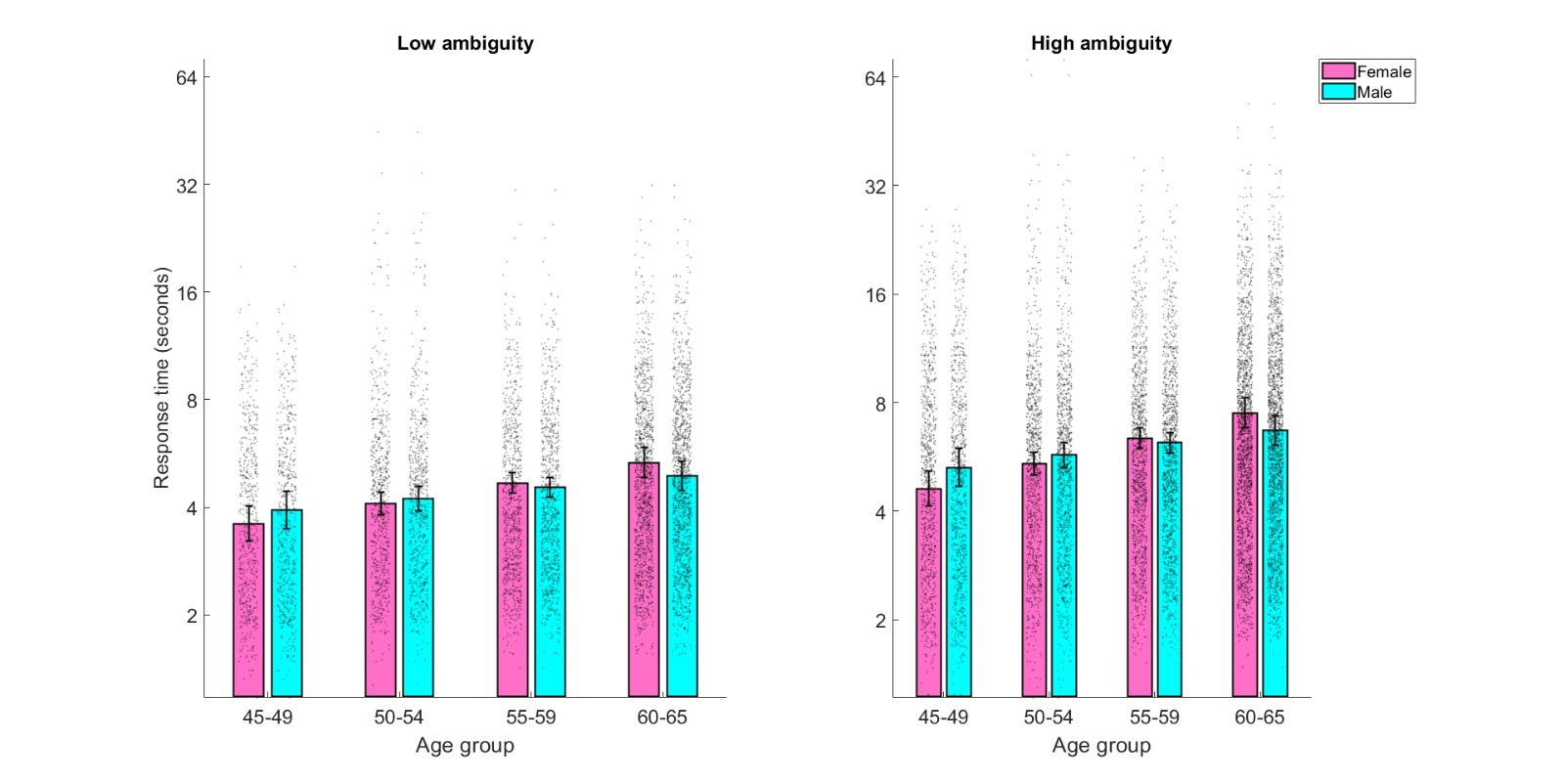
